# The Plasma Glycome of Women With PCOS is Different From Healthy Controls

**DOI:** 10.1101/2025.03.14.25323948

**Authors:** Madison Holman, Sophie Jie Li, Mary M Ahern, L Renee Ruhaak, Siddika Karakas, Sridevi Krishnan

**Author notes:** Corresponding author: Sridevi Krishnan, PhD, Thomas W Keating Bioresearch Building 1657 E Helen St Tucson AZ 85721. Co-first authors listed alphabetically (Madison Holman, Sophie Jie Li).

## Abstract

**Objective/Background:** While PCOS research has extensively explored genomic, transcriptomic, proteomic, and metabolomic milieus, our study examines the plasma glycome, comparing women with PCOS to age-matched healthy controls.

**Methods:** In this observational study, an n = 47 women with PCOS were screened and enrolled at the UC Davis Health campus, the comparator group constituted an n = 25 age-matched healthy women. During a study visit, body composition was measured using a bioelectrical impedance scale, and fasted plasma samples were obtained to measure glucose, insulin, circulating lipids and leptin, among other parameters, in all groups. In addition, in the PCOS group, circulating androgens and other endocrine hormones were measured. The plasma glycome was measured using a UHPLC-MS protocol.

**Results:** As expected, women with PCOS had more body weight (p<0.01), body fat (p = 0.004), fasting leptin (p=0.01), insulin (p = 0.003) and glucose (p = 0.004). The plasma glycome milieu displayed increased tetraantennary (glycans with 4 branches: p = 0.05) and reduced hybrid-type glycans (p = 0.019) in women with PCOS compared to healthy controls. Logistic regression models predicting PCOS vs control binary outcomes indicated a higher tetraantennary and lower hybrid glycan profile to represent women with PCOS more than controls both with (AUCROC = 0.94) and without body weight (AUCROC = 0.84). Further, in women with PCOS, total testosterone was positively associated with tetra-antennary glycans (r = 0.322, p = 0.029).

**Conclusions:** Highly branched glycans, that have been shown to be elevated in pro-inflammatory and metabolic disease states, are also elevated in PCOS. However, the link between circulating androgens and protein glycosylation in women remains unknown, and future investigations should focus on this.

## Introduction

Polycystic ovary syndrome (PCOS) is the most common endocrine disorder affecting one out of 10 women of reproductive-age[1]. PCOS is a complex disorder: its reproductive manifestations include oligomenorrhea/oligo-ovulation or amenorrhea and androgen excess; its metabolic manifestation is insulin resistance, and these symptoms vary on a spectrum[2]. There is currently no established root cause for the imbalance of hormones, and subsequently, no cure and treatment plans primarily address symptoms[2]. A fascinating aspect of this syndrome is that relatively small amounts of weight loss (5-7%) and /or treatment of insulin resistance improves reproductive function in PCOS, indicating link between the two facets of this disorder.

Research employing “omic” technologies have contributed a great deal to understanding of PCOS providing valuable insights into the molecular pathways involved[3]. PCOS is strongly heritable[4] and polygenic[5] and genome-wide association studies have been instrumental in identifying susceptibility loci and candidate genes for PCOS, such as *LHCGR*, *ERBB4*, *FSHR*, and more [6–8]. Mutations in the FSHR/LHCGR and INSR loci have been identified in women with PCOS [9]. Owing to its polygenic nature, genetic variants associated with insulin signaling, glucose homeostasis, and metabolic abnormalities in PCOS patients have been observed [10, 11]. In addition, epigenetic changes in genes controlling skeletal muscle metabolism have been observed in women with PCOS, potentially contributing to the metabolic disturbances seen in these women [11]. To add further complexity, the involvement of extraovarian androgen actions mediated by genes such as ERBB4/HER4, YAP1, and FSHB in PCOS suggests that it is not solely an ovarian disorder [12, 13]. Also, impaired insulin-stimulated glucose disposal in PCOS patients has been linked to reduced expression of nuclear-encoded genes involved in mitochondrial oxidative metabolism [14]. Finally, studies have shown that PCOS is associated with increased risk for cardiovascular diseases, metabolic syndrome, dyslipidemia, type 2 diabetes, and impaired fertility [15]. These further support the polygenic nature of the etiology of PCOS.

Follicular fluid lipidomic profiles in different phenotypes of PCOS identified increased total ceramides, total fatty free acids, decreased lysophosphatidylglycerol (LPG,18:0, LPG,18:1, and LPG,18:2), increase triglycerides, phosphatidylethanolamines & phosphatidylinositols in women with PCOS than controls[16, 17]. Higher circulating concentration of ceramides (Cer 36:1;2 - (Cer(d18:1/18:0) - N octadecanoyl-sphing-4-ene)) have been linked with insulin resistance, a common symptom in PCOS [16]. Gut microbiota and metabolomic studies have identified certain bacterial species that produce changes in metabolism independent of diet [18, 19]. For example, some intestinal flora components like Clostridiales, Bacteroidetes, and Porphyromonadaceae differ in people with PCOS [20–22]. Trimethylamine N-oxide (TMAO), a byproduct of red-meat consumption is associated with hyperandrogenism, and is considered a possible metabolite involved in the pathogenesis of PCOS [20].

The N-glycome has, in recent years, become critical to understand in the role it plays in metabolic health[23]. The N-glycome is the set of all sugars/glycans/oligosaccharides added to proteins in posttranslational or co-translational processes[24]. N-glycans have both structural and functional roles, disruption of which can disrupt homeostasis. In particular, sialylation (the addition of the 9-carbon sialic acid sugars to the protein), fucosylation (addition of 5-carbon fucose), and branching (increased bisecting GlcNAcs and/or antennae) have been found to have profound influences in immune and inflammatory diseases, cancer progression, and metabolic disease onset [25–28]. Both due to their involvement in disease processes as well as being a result of disease processes[23], the N-glycome could hold pertinent information that helps further our comprehension of poorly understood diseases, such as PCOS. Hence, in the current manuscript, we compare the plasma glycome of women with PCOS against age-matched healthy controls, to explore differences that may be observed. Since the influence of sex-steroid hormones (estrogens and androgens) on the glycosylation process is poorly understood[29], it is difficult to ascertain whether these are causal or reactive changes. However, this is the first step in furthering our understanding of differences, if any, that do exist.

## Methods

This report includes a cross-sectional comparison of plasma glycome in women with PCOS compared to a control group of women who are healthy.

### Participants

This study utilized archived samples and data from previously conducted intervention studies[30–32] all approved by the Institutional Review Board of the University of California, Davis. The authors had access only to deidentified data and plasma samples. Participants for the original studies were recruited between September 2007 and February 2010, with written informed consent obtained prior to inclusion. For the current investigation, women aged 20–45 years with a BMI of 25–45 kg/m² were included in the control group. The PCOS group met the same demographic and anthropometric criteria but additionally fulfilled the NIH diagnostic criteria for PCOS, requiring ovarian dysfunction (amenorrhea: >6 months without menstruation, or oligomenorrhea: <6 periods per year), clinical or biochemical evidence of hyperandrogenemia (total testosterone >54 ng/dL or free testosterone >9.2 pg/mL), and absence of confounding conditions (e.g., Cushing’s disease, 21-hydroxylase deficiency, or prolactinoma). Exclusion criteria included recent use (within two months) of oral contraceptives, anti-androgenic medications, insulin sensitizers, d-chiro inositol, or lipid-lowering drugs, as well as diabetes, untreated thyroid disorders, systemic illnesses (renal, hepatic, or gastrointestinal diseases), smoking, or consuming more than two alcoholic drinks per week.

Data collection occurred at the Clinical and Translational Science Center Clinical Research Center (CCRC) at the University of California, Davis. Menstruating women were tested during the follicular phase of their cycle. All participants provided an overnight fasted blood sample via venipuncture, which was processed, aliquoted, and stored at –80°C until analysis.

### Anthropometric data

Weight was measured in light clothing using the Tanita BWB800-P Digital Medical Scale. Body composition was determined using bioelectrical impedance 13 (Biostat Co., Isle of Man, UK)[33]

### Biochemical measurements

Fasting glucose was measured using YSI 2300 STAT Plus Glucose & Lactate Analyzer (YSI Life Sciences, Yellow Springs, OH), with a coefficient of variation (CV) of 1%. Fasting Insulin was measured by RIA (Milipore, St. Charles, MO) with a CV of 8.2%. Hepatic insulin resistance was assessed by calculating Homeostasis Model Assessment (HOMA: [(fasting insulin (μU/ml) × fasting glucose (mg/dl))/405. Triglyceride, cholesterol and HDL-cholesterol were measured using Poly-Chem System Analyzer (Cortlandt Manor, NY) with CVs of 3.5%, 4% and 3.6%, respectively. Leptin and adiponectin were measured using RIA (Millipore, St. Charles, MO) with CVs of 4.3% and 6.5%. hs-CRP was measured using a highly sensitive (hs) latex-enhanced immunonephelometric assay with a CV <5%. Total testosterone, sex hormone binding globulin (SHBG) and DHEAS were measured by RIA (Diagnostic Systems Laboratories, Webster, TX) with CVs of 8.3%, 4.4% and 9.6%, respectively.

### N-glycan preparation

All glycome analyses were completed between 2012 and 2013 in samples that were stored at -80 Deg C. N-glycans were released from plasma as previously described[34, 35]. In brief, N-glycans were prepared from 50 µL of human plasma after protein denaturation with 50 μL of 200 mmol/L ammonium bicarbonate (Sigma-Aldrich) solution with 10 mmol/L dithiothreitol (Promega). Two µL of PNGase F (New England Biolabs) was added to the samples and glycans were enzymatically released in a CEM microwave, followed by purification and enrichment by solid phase extraction using PGC on a 96-well plate. Released N-glycans were dried in vacuo prior to analysis.

#### nHPLC-chipTOF-MS analysis

The dry N-glycan sample was reconstituted in 225 μL of water and transferred to an injection vial for analysis; 1 μL of sample was used for injection. N-Glycans were analyzed using an Agilent nanoHPLC-chip-TOF-MS, in which a PGC chip II (Agilent Technologies, Santa Clara, CA) packed with porous graphitized carbon was used. Upon injection, the sample was loaded onto the enrichment column and subsequently separated on the analytical column using a gradient of 3 % acetonitrile (ACN) with 0.5 % formic acid (FA) (solvent A) to 90 % ACN with 0.5 % FA (solvent B). The mass spectrometer was operated in positive mode.

#### Data analysis

Data analysis was performed using Masshunter® qualitative analysis according to Hua et al.[36] with modifications. Data were loaded into Masshunter® qualitative analysis, and glycan features were identified and integrated using the Molecular Feature Extractor algorithm. A retrosynthetic theoretical glycan library containing 331 possible N-glycan compositions was used for glycan identification[37]. To increase biological relevance, similar types of glycans were grouped and summations calculated. For instance, all glycans that had a single sialic acid were summed into a category labeled monosialylated glycans. This was repeated for sialylations and fucosylations. In addition, glycans were classified and summed based on their structure (see **Supplemental table 1**).

#### Statistical analysis

Data analyses were done using R[38] and JMP Pro 18.1 (SAS Institute, Cary NC). Data missingness was evaluated using Amelia package in R[39]. The missingness map (**Supplemental Fig 1**) indicated a 12% of all data (and 28% of clinical data) was missing. Since a significant portion of the clinical data were missing, data imputation was not conducted and final data analysis included parameters that did not have missing data, and used only complete case data. Data normality was evaluated by Q-Q plots, Shapiro-Wilk tests (11% of data was normally distributed) and Kolmogorov-Smirnov tests (0% of data was normally distributed). Data were either normalized for further analyses (Johnson transformations), or non-parametric tests were used where available.

Given the wide and short dataset (n << parameters), a partial least squares discriminant analysis (PLS-DA) was used for dimension reduction with all individual glycans and summated glycan sub-types to identify their ability to discriminate between the PCOS vs control groups. For the PLS-DA, normalized data were used in a NIPALS algorithm, using leave-one-out cross validation (since the dataset was small and this approach would not be computationally intensive) and a VIP (variable importance in projection) score of >1 used as a cut-off that deems a certain parameter strong enough to discriminate between PCOS and controls. Going forward, only the glycomic parameters with a VIP score>1 were used in analyses, unless otherwise specified. Wilcoxon tests were used to determine whether there were significant differences in selected glycomic variables and clinical parameters between women with PCOS and healthy controls; p-values of <0.05 were considered significant. Spearman’s rho tests were used to evaluate correlations between selected glycomic and clinical parameters for the PCOS and control groups. Logistic regression modelling was used to evaluate the predictive ability of all summed glycans to classify PCOS and healthy controls. In addition, these models were also used to evaluate the effect of addition or removal of body weight and body composition parameters on the predictive ability of glycans in classifying PCOS and healthy controls. The AUROC (area under the receiver operator curve) and VIF (variable inflation factor) were used to evaluate model and parameter fit.

## Results

### Clinical characteristics of study participants

A total of 72 women were included in this study, 47 with PCOS and 25 healthy controls. Differences in demographic and clinical outcomes can be seen in **Table 1**. Women with PCOS had significantly higher BMI (35.5 ± 7.9 kg/m^2^) compared to healthy controls (25.7 ± 3.5 kg/m^2^), but were not different in age (32.0 ± 6.3 vs. 30.1 ± 8.1). Women with PCOS had significantly higher fat mass (44.0 ± 16.3 kg) as well as higher lean mass (53.2 ± 5.5 kg) than control fat mass (27.5 ± 4.8 kg) and lean mass (46.0 ± 4.5). Healthy women had significantly lower levels of CRP (1.8 ± 1.3 mg/L) than levels of CRP in women with PCOS (6.0 ± 7.1 mg/L). Women with PCOS had higher fasting insulin (PCOS: 20.4 ± 9.8 mIU/mL vs. Control: 11.9 ± 3.5 mIU/mL), glucose (PCOS: 98.1 ± 12.3 mg/dL vs. Control: 90.1 ± 7.7 mg/dL), leptin (PCOS: 24.9 ± 12.9 ng/mL vs. Control: 15.2 ± 6.1 ng/mL), and adiponectin levels (PCOS: 9.6 ± 5.6 ng/mL vs. Control: 12.9 ± 5.4 ng/mL). HDL-c was lower (40.2 ± 9.3 mg/dL) in women with PCOS while Triglycerides were higher (107.5 ± 51.0 mg/dL). No significant difference was observed in total cholesterol or triglycerides between the two groups.

**Table 1:**
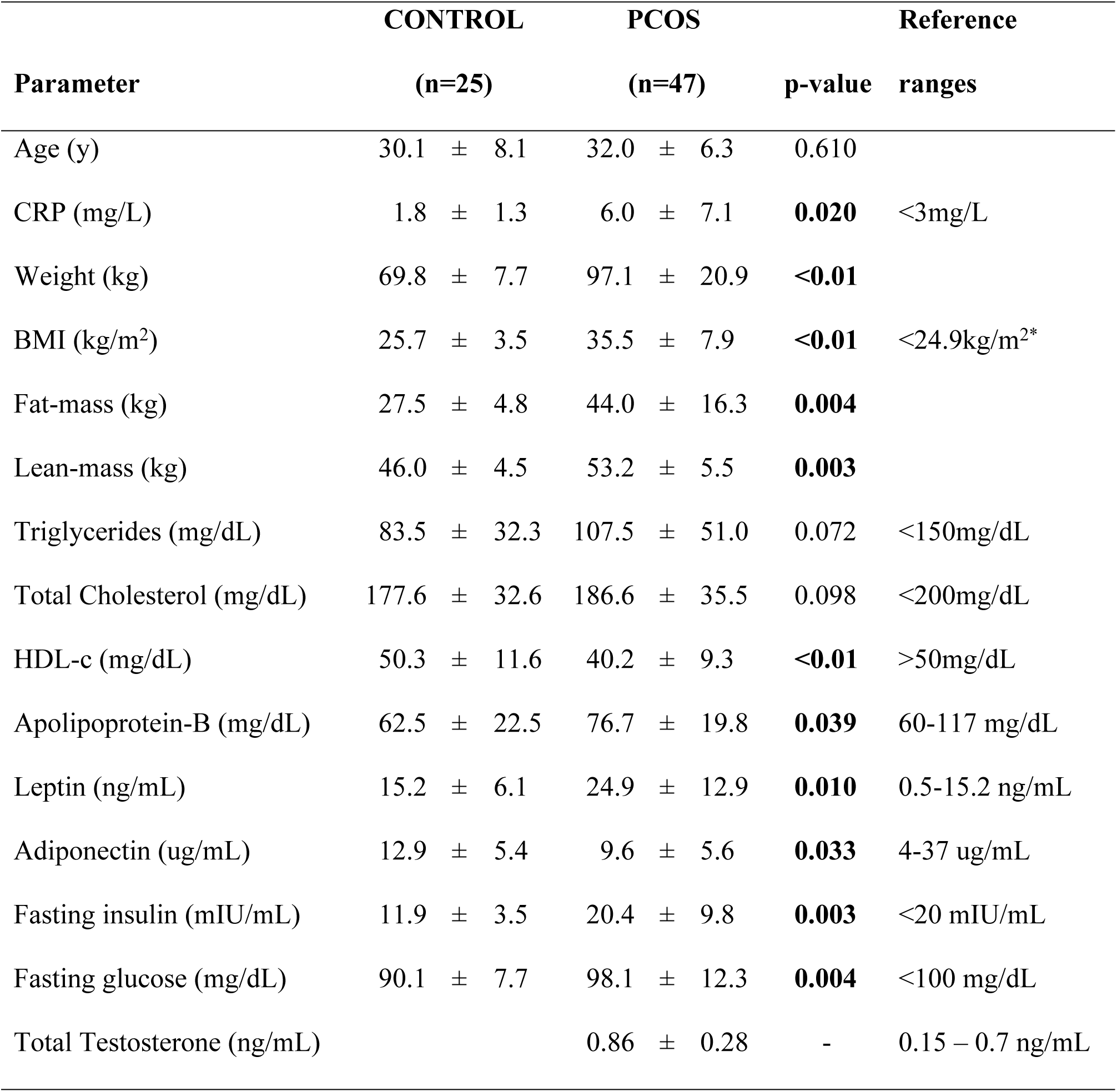
Clinical Data of the Control and PCOS groups. Data is presented in mean ± standard deviation. BMI, body mass index. CRP, C-reactive protein. HDL-c, high-density lipoprotein cholesterol. P-values reported are for the relationship between PCOS and healthy groups. *BMI of <25 is for normal weight individual

#### PLS-DA for dimension reduction

The PLS-DA model that provided the best discrimination between PCOS and control group had 2 factors with a root mean PRESS value of 0.986, an R^2^X of 0.10 and R^2^Y of 0.77, and Q2 of 0.02. This suggests that 10% of the variation between PCOS and controls were explained by ∼77% of variation in the plasma glycome. Since the model only explains 10% of the variation between PCOS and controls, and its discriminatory ability is quite low (Q2 < 0.2) , a follow-up PLS-DA model using variables with VIP score > 1 in this iteration as independent parameters, predicting PCOS and controls was evaluated. This model also had 2 factors, with a root mean PRESS of 0.791, and R^2^X of 0.2 (20%) and R^2^Y of 0.856 (85.6%) with a Q2 value of 0.37. Only VIP variables with score > 1 present in both PLS-DA models were used in all further analyses, unless otherwise specified.

#### Individual glycan differences in healthy controls and women with PCOS

VIP score >1 variables from the PLS-DA model above were used to identify glycomic profile differences between women with PCOS and healthy controls, and are depicted in Figure 2. Women with PCOS exhibited significantly higher concentrations of fMan4GlcNAc4NeuAc1 (p < 0.001), Man5GlcNAc4 (p < 0.001), Man6GlcNAc3 (p = 0.004), Man6GlcNAc3NeuAc1 (p = 0.008), Man4GlcNAc3 (p = 0.008), Man4GlcNAc4 (p = 0.011), Man5GlcNAc3NeuAc1 (p = 0.016). Conversely, women with PCOS exhibited significantly lower mean levels of Man6GlcNAc5Fuc1NeuAc2 (p = 0.018) and Man3GlcNAc4Fuc1 (p = 0.044) compared to healthy women.

**Figure 1:**
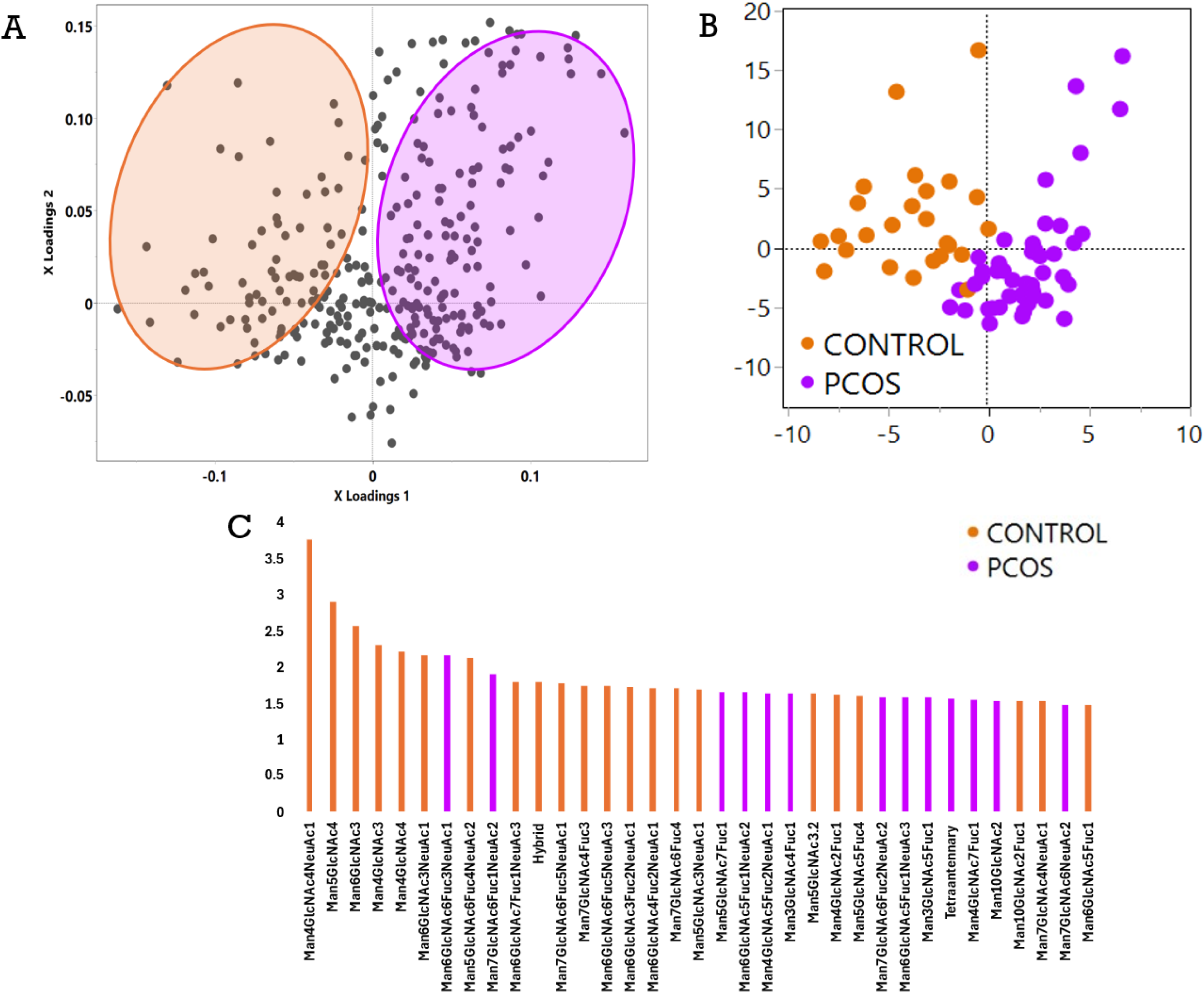
A partial least squares discriminant analysis depicting the ability of the plasma glycome in differentiating between women with PCOS (purple) and healthy controls (orange). A depicts the loadings plot where each grey circle is one glycan species. B depicts the scores plot, showing the discriminatory ability of the PLSDA model to differentiate between women with PCOS and healthy controls. C is the VIP (variable importance in projection) plot where the Y-axis shows the VIP score, and the X axis shows the individual or summed glycans that are the strongest discriminators between women with PCOS and healthy controls.

**Figure 2:**
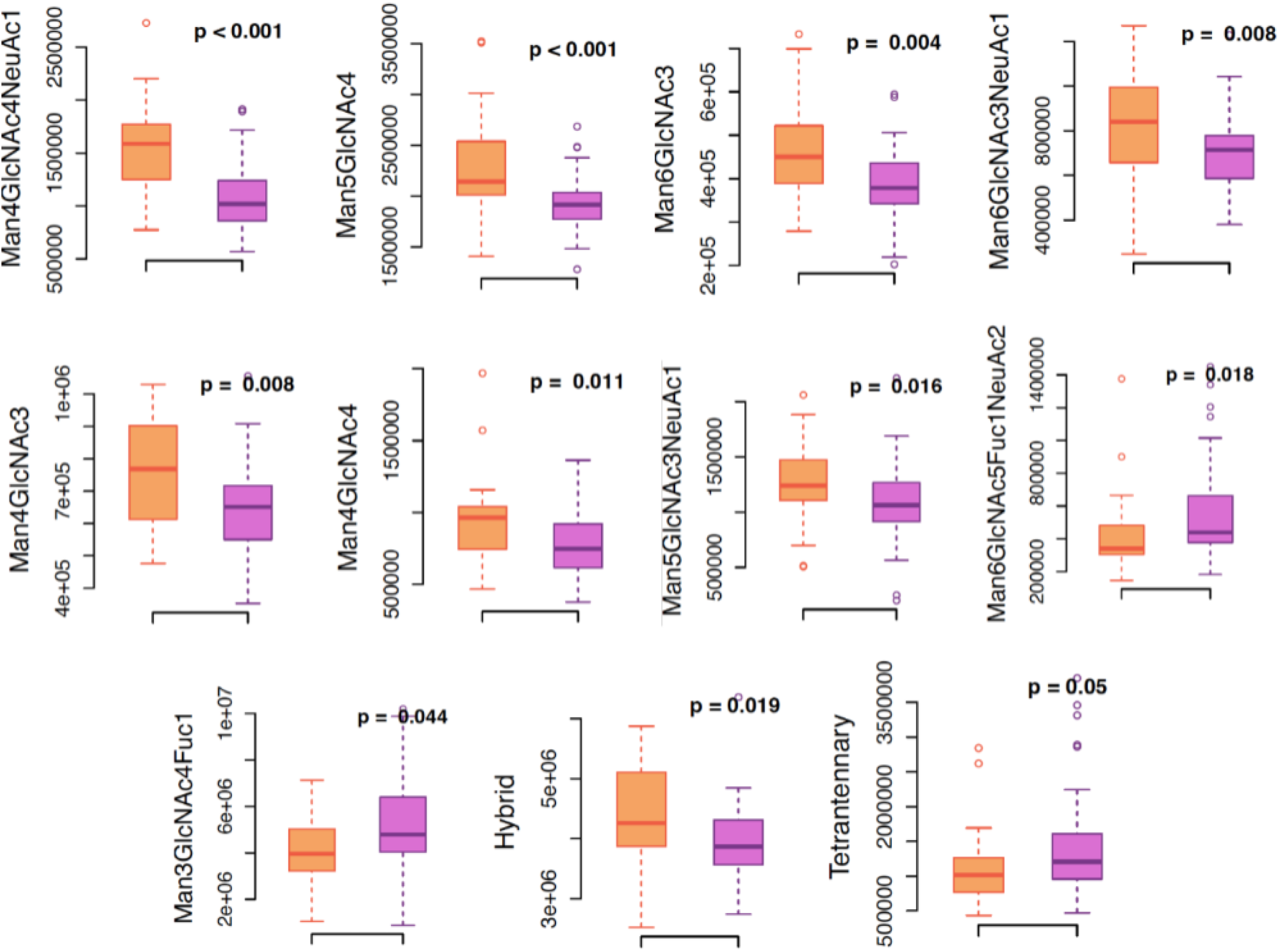
Differences between women with PCOS (depicted in purple) and healthy controls (orange) in glycomic features. Among several glycans, while women with PCOS had lower hybrid glycans compared to healthy controls, they had higher tetra-antennary glycans in plasma.

#### Glycan summation differences in healthy controls and patients with PCOS

In Figure 3, women with PCOS had a significantly higher concentration of hybrid glycans (p = 0.019) than healthy controls. However, women with PCOS had lower tetra-antennary glycans (p = 0.05) compared to healthy controls.

**Figure 3.**
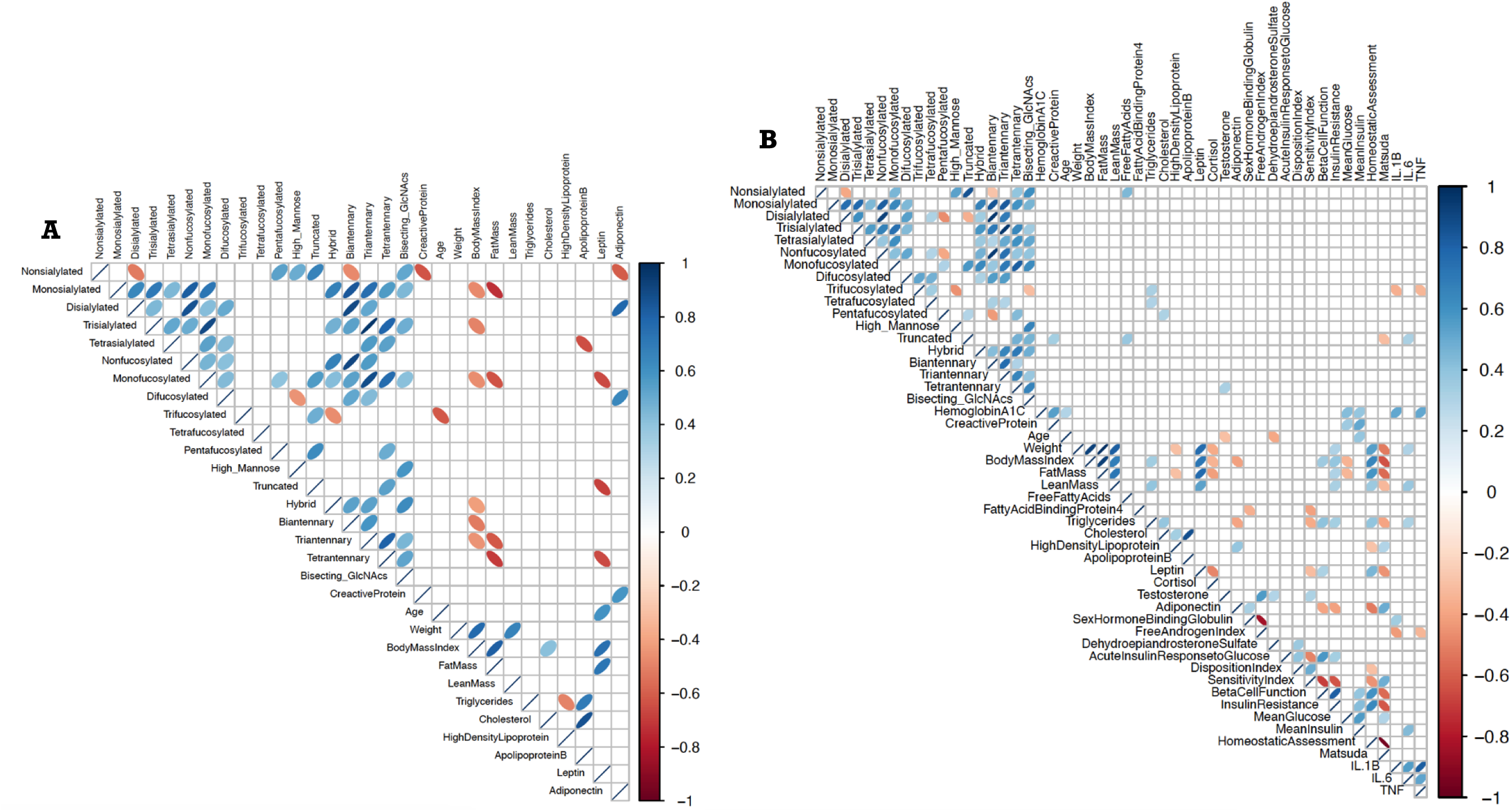
Correlogram of glycomic, anthropometric, clinical and metabolic features of healthy controls (n = 25) [**A]** compared to women with PCOS (n = 47) [**B].** Significant positive associations (based on Spearman’s rho) are depicted by blue ellipses, and inverse associations by orange/red ellipses, light vs dark shades of same color indicate weak vs strong associations, thick vs thin ellipses (if it almost resembles a line) represents the wider vs tighter 95% confidence interval around the scatter area. Women in the healthy control group display positive relationships between leptin and oligomannose rich glycans as well as sialylated and fucosylated glycans with adiponectin, while these relationships are either weaker in women with PCOS or non-existent.

#### Correlation analysis

The PCOS group had more endocrine and clinical information than the control group, resulting in a larger correlogram. It is of interest to note that testosterone was positively associated with tetra-antennary glycans (r = 0.322, p = 0.029) in PCOS (Fig 3). Curiously, in the control group leptin (and fat-mass) was inversely associated with tetra-antennary (r = -0.650, p = 0.026), truncated (r = -0.685, p = 0.017) and mono-fucosylated glycans (r = -0.657, p = 0.024). Furthermore, adiponectin was inversely associated with non-sialylated glycans (r = -0.608, p = 0.040), and positively associated with di-sialylated (r = 0.783, p = 0.004) and di-fucosylated (r = 0.643, p = 0.028) glycans. In the PCOS group, however, no significant associations were observed between leptin/adiponectin and the plasma glycome.

#### Logistic Regression analysis

To identify whether the glycome differences we observe between PCOS, and control groups of women is primarily due to their body weight, we used logistic regression models. These models were made with glycomic variables as independent predictors with and without the inclusion of body weight ad body fat to predict PCOS and control classes. Since the goal was to represent the whole glycome in this model, all glycan summations were used. Figure 4 represents the outcomes of these models. When just the glycan summations were used to predict classes – PCOS and controls, hybrid, tetra-antennary and non-sialylated glycans were identified as significant predictors. However, the non-sialylated glycans had a VIF greater than 5[40], suggesting high multicollinearity with other independent variables, and making the identified relationship between non-sialylated glycans and outcome groups (PCOS vs control) less reliable. The addition of body weight to the model resulted in the retention of tetra-antennary and hybrid glycans as significant predictors. This did not change when fat mass was used instead of body weight. In addition, when fat mass was used non-sialylated, truncated, oligo-mannose and fucosylated glycans became significant predictors as well, however, non-sialylated, truncated and high-mannose glycans again had very high VIF, suggesting unreliable model estimates. In all models, hybrid glycans were positive predictors of the control group whereas tetra-antennary glycans were positive predictors of the PCOS group.

**Figure 4.**
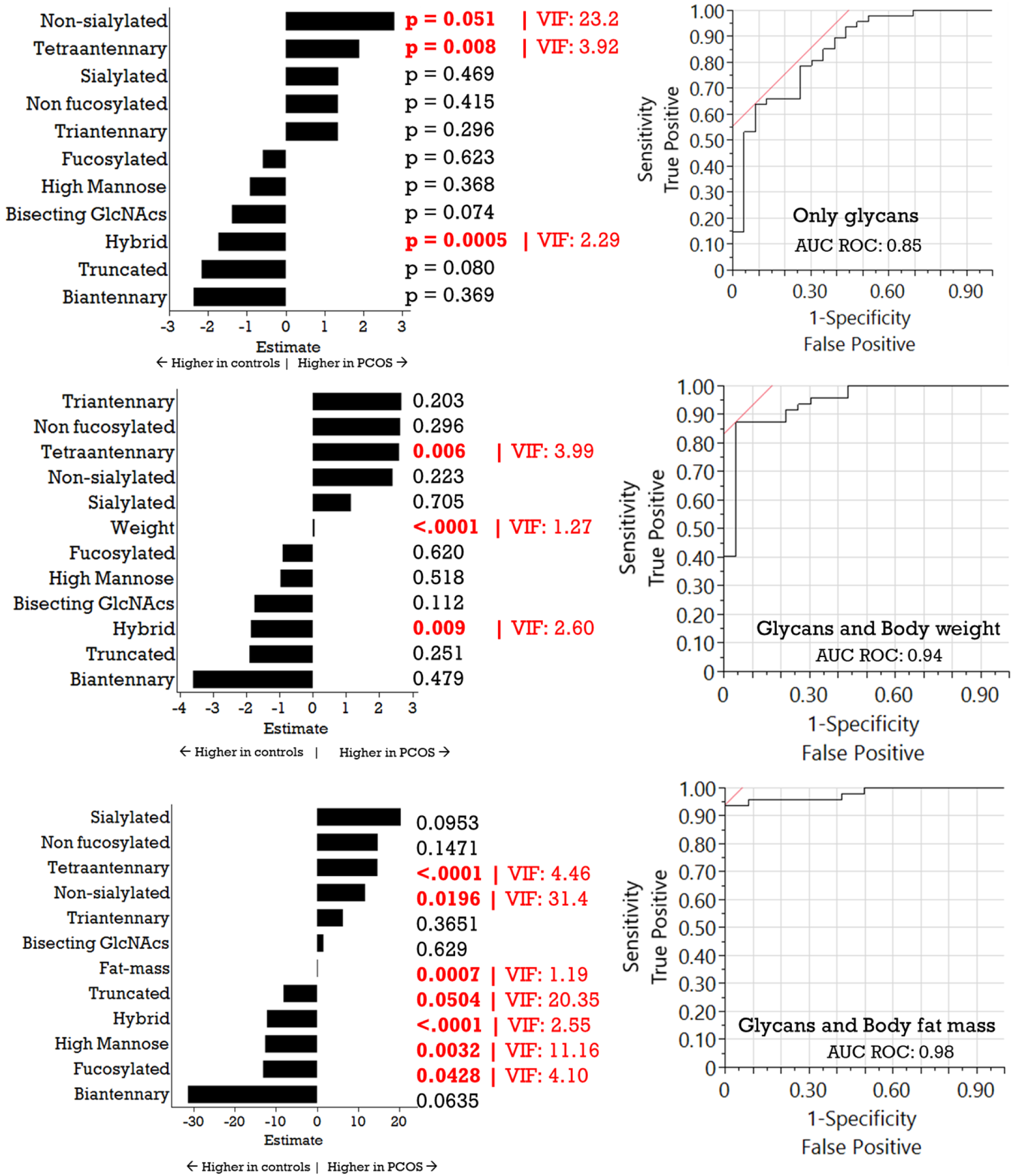
Logistic regression analysis to evaluate the role of addition of body weight and adiposity to models classifying healthy controls from women with PCOS. ***Top***: Model estimates (left), their corresponding p-values, the variable inflation factor (VIF) and the AUC ROC (right) showing summated glycans and their ability to classify women with PCOS from healthy controls. ***Middle***: Model outcomes of model with the addition of body weight. ***Bottom***: Model outcomes with the addition of body fat. Hybrid and tetra-antennary glycans are consistent classifying glycans across all the models.

## Discussion

In this cross-sectional comparative study of the plasma glycome of PCOS women against healthy controls, tetraantenary glycans were found elevated in PCOS, while hybrid glycans were lower. While the higher branched glycans have been shown to be associated with metabolic diseases like insulin resistance and cardiovascular disease, they have also been associated with obesity. Our logistic regression analyses, however, suggests that in women with PCOS, the increase in tetraantenary glycans and lower abundance of hybrid glycans could be independent of body weight. The inverse associations observed between leptin/fat mass and tetra-antennary glycans in the control group were not seen in women with PCOS, which, speculatively, could hint at disruptions of healthy biological relationships. Further, we found significant associations between highly branched glycans and testosterone, one of the primary androgens involved in precipitating the ovarian disruptions observed in PCOS. This suggests that further research is necessary to explore the possibility of the ratio of branched to hybrid glycans as detection biomarkers for certain phenotypes of PCOS.

Tetraantennary glycans, characterized by their branched structure with four antennae, play a significant role in various biological processes, including metabolic health. The presence of tetraantennary structures has been associated with increased inflammation[41] and the presence of metabolic conditions, including type 2 diabetes and cardiovascular diseases[42]. There have been significantly higher levels of tetra-antennary glycans found in plasma in women with endometriosis[43], ovarian cancer and even higher levels as the disease progresses compared to healthy individuals[44–46]. These changes in glycosylation patterns may serve as potential biomarkers for the diagnosis and monitoring of ovarian cancer[44, 45]. One of the critical mechanisms by which tetraantennary glycans influence metabolic health is through their role in modulating inflammation[42]. In addition, tetraantennary glycans are implicated in the regulation of lipid metabolism. Chan et al. highlighted that specific glycan structures can influence the metabolism of very-low-density lipoproteins (VLDL) and proprotein convertase subtilisin/kexin type 9 (PCSK9), both of which are crucial in lipid homeostasis[47]. The interaction between these glycans and lipoproteins can affect the clearance of triglyceride-rich lipoproteins from circulation, thus playing a role in the development of dyslipidemia, which is often observed in metabolic syndrome, obesity and PCOS[47, 48].

The synthesis of glycans is mediated by a series of glycosyltransferases that add specific sugar residues to growing glycan chains. When more branched glycans, such as tetraantennary or highly branched complex glycans, are synthesized, they may compete for the same substrates and enzymes that are responsible for producing hybrid glycans[49]. The presence of additional sialic acid residues and branching can enhance the retention of certain glycans, potentially leading to the exclusion of hybrid glycans from the glycan pool[50]. Furthermore, the presence of branched structures can enhance the binding affinity of certain glycan-binding proteins, indicating that the glycan structure can influence enzymatic pathways and potentially reduce the synthesis of hybrid-type glycans[51]. Via such mechanisms, a milieu that promotes more tetraantennary glycans could influence the glycome to have less hybrid type glycans. Hybrid type glycans play critical roles in fertility. Reductions in hybrid-type glycans in oocytes has been shown to reduce its ability to be fertilized[52]. Furthermore, oocytes lacking hybrid type glycans also do not mature past embryonic day 9.5 during fertilization[53, 54]. Thus, a systemic circulatory milieu that has more tetra- antennary and lower hybrid glycans could be involved in and reflect the metabolic and fertility disruptions seen in women with PCOS.

In the current study, we found an association between tetraantennary glycans and testosterone in the PCOS group. The relationship between higher testosterone levels in women with Polycystic Ovary Syndrome (PCOS) and metabolic dysregulation is well-documented in the literature. Elevated testosterone, a hallmark of PCOS, is associated with various metabolic disturbances, including insulin resistance, dyslipidemia, and an increased risk of type 2 diabetes and cardiovascular disease. higher levels of free testosterone are correlated with increased insulin resistance, as measured by the Homeostasis Model Assessment of Insulin Resistance (HOMA-IR)[55, 56]. Free testosterone levels were significantly associated with insulin resistance in women with PCOS, indicating that hyperandrogenism may exacerbate metabolic dysregulation[56].

Insulin resistance has been linked to changes in the glycosylation patterns of plasma proteins in women with Gestational Diabetes Mellitus[57]. Specifically, insulin resistance is associated with a decrease in low-branched glycans and an increase in high-branched, like tetraantennary, glycans on plasma proteins[57]. This shift in glycosylation patterns is typically observed in type 2 diabetes, which can be a consequence of insulin resistance. Glycosylation, in turn, can also influence insulin sensitivity and signaling, as shown by the lowered glucose transporting capacity of hypoglycosylation of GLUT4[58].

In men, low testosterone levels are associated with the metabolic syndrome, type 2 diabetes, and obesity, and that testosterone replacement therapy can improve glycemic control and insulin resistance[59–61]. The underlying mechanisms may involve the influence of testosterone on the glycosylation of key proteins involved in glucose and lipid metabolism. And these are yet to be investigated. Furthermore, the effect of testosterone on protein glycosylation in women is unknown. Future research will have to investigate the relationship between circulating estrogens, androgens and protein glycosylation, to establish a baseline understanding of the “endocrine-glycome” axis.

### Limitations, Strengths, Conclusions and Future Directions

The current study is a cross-sectional investigation, and only able to report observational differences between healthy vs women with PCOS, and associated analyte relationships. However, no study till date has evaluated the plasma glycome in women with PCOS, thus marking the novelty of this manuscript. A higher tetraantennary, and lower hybrid-type glycan plasma profile in women with PCOS suggests the presence of a pro-inflammatory metabolic disease-like milieu. While this is not surprising, it lends itself to future investigations that can identify if these glycans can be used to better detect PCOS phenotypes, which can then inform appropriate therapeutic approaches.

## Data Availability

All data will be made available on our github page upon manuscript acceptance - https://github.com/Glyconutlab.

## Acknowledgements and author contributions

SKs and RLR collected (and oversaw the collection of) clinical and glycomic data, MH, SJL, MMA and SKn conducted data analyses, MH and SJL wrote the initial manuscript draft, SKn and SKs had significant editorial input, all authors helped complete writing the manuscript.

